# Evidence for bilingualism as a cognitive reserve factor in biomarker-confirmed Alzheimer’s disease

**DOI:** 10.64898/2026.03.31.26349879

**Authors:** Stephanie M. Grasso, Wenfu Bao, Sonia Karin Marqués-Kiderle, Natalia Casart Muñoz, Marco Calabria, Isabel Sala, Maria Belén Sánchez-Saudinós, Elena Vera-Campuzano, Judit Selma-González, Laura Videla, Lídia Vaqué-Alcázar, Alexandre Bejanin, Jesús García-Castro, Íñigo Rodríguez-Baz, Nuole Zhu, Javier Arranz, Lucia Maure-Blesa, Sara Rubio-Guerra, Isabel Barroeta, Ignacio Illan-Gala, Maria Carmona-Iragui, Olivia Belbin, Daniel Alcolea, Juan Fortea, Alberto Lleó, Miguel Ángel Santos Santos

## Abstract

**INTRODUCTION:** Bilingualism is a proposed cognitive reserve factor that delays symptom onset in Alzheimer’s disease (AD), though current evidence lacks biomarker confirmation. This retrospective study examined bilingualism’s association with symptom onset across AD clinical stages, including biomarker-confirmed cases.

**METHODS:** Participants from the Sant Pau Memory Unit spanning amnestic mild cognitive impairment (MCI), amnestic dementia, and biomarker-confirmed AD were analyzed, with balanced representation of active and passive Spanish-Catalan bilinguals. Linear regression models evaluated associations between bilingualism and reported age at symptom onset, controlling for education, sex, and disease severity.

**RESULTS:** Active bilingualism was associated with delayed symptom onset in amnestic MCI (2.21 years), amnestic dementia (1.42 years), and biomarker-confirmed AD (1.45 years; *p*s < .05). Higher education was associated with earlier onset, likely representing healthcare seeking behavior.

**DISCUSSION:** Bilingualism protects against earlier symptom manifestation in MCI and AD, supporting bilingualism as a contributor to cognitive reserve.

## 1. Background

Alzheimer’s disease (AD) is the most common cause of dementia, accounting for approximately 60–70% of cases worldwide, and its prevalence continues to rise with population aging [1–3]. Clinically, AD is characterized by progressive impairment in memory, language, executive functioning, and behavior, while neuropathologically it is defined by extracellular amyloid β (Aβ) plaques and intracellular neurofibrillary tangles, with early involvement of the hippocampus and associative cortices [1]. Despite shared biological features, individuals with comparable levels of AD pathology often differ substantially in the age at which clinical symptoms emerge, suggesting that modifying factors influence the translation of neuropathology into clinical impairment.

The concept of cognitive reserve has been proposed to explain this variability, positing that lifetime cognitive enrichment enables individuals to better cope with brain pathology and delay the onset of clinical symptoms [4]. Within the context of AD, contemporary frameworks further distinguish between resilience, defined as better-than-expected cognitive performance given the level of pathology, and resistance, referring to reduced accumulation of pathology itself [5–7]. These constructs are complementary and provide a useful lens for understanding how environmental and lifestyle factors modulate both the biological burden of disease and its clinical expression [4,8].

Bilingualism has emerged as one life experience that contributes to cognitive reserve. The regular use of two languages requires sustained engagement of executive control processes, including attentional regulation, inhibition, and switching, which are repeatedly recruited during language selection and monitoring [9–11]. Neuroimaging studies in individuals with mild cognitive impairment (MCI) and dementia indicate that bilingualism modifies the relationship between neuropathology and cognition through two main mechanisms. Bilingual participants often show greater cortical atrophy or hypometabolism in AD-vulnerable regions despite comparable cognitive performance, consistent with cognitive resilience, while some studies also report relative preservation of subcortical structures or slower atrophy rates, suggesting region-specific resistance mechanisms [12–14]. Together, these findings support a dual contribution of bilingualism to resilience and resistance processes that may delay the clinical expression of neurodegenerative pathology.

At the clinical level, multiple retrospective studies and meta-analyses report that bilingual individuals experience a delayed onset of dementia symptoms, including AD, by approximately four to five years compared with monolinguals [15–23]. Importantly, this delay has often been observed despite comparable or greater levels of brain atrophy or hypometabolism at diagnosis, supporting a resilience-based interpretation [12]. However, much of this literature relies on syndromic clinical diagnoses without biomarker confirmation, which may lead to etiological misclassification and limits mechanistic inference [24,25]. In addition, heterogeneity in how bilingualism is operationalized across studies and the potential presence of confounding variables (e.g., education, biological sex) complicate cross-study comparisons and may partially account for inconsistent findings [26].

Recent work from our group has begun to address these limitations by integrating biomarker data within a resilience–resistance framework [27]. In a cohort of Spanish-Catalan bilinguals with biomarker-confirmed AD, we demonstrated that active bilinguals, defined by the regular use of both languages, outperformed passive bilinguals across executive, language, and visuospatial domains despite equivalent biological disease stage, providing evidence of cognitive resilience. Moreover, active bilinguals showed more favorable cerebrospinal fluid (CSF) and plasma biomarker profiles related to amyloid burden and neuroinflammation, suggesting concurrent resistance to AD-related pathophysiology.

In the sociolinguistic context of Barcelona, categorizing bilingualism as active versus passive is particularly appropriate. Catalan-dominant individuals nearly universally acquire and use Spanish, whereas Spanish-dominant individuals vary in their active use of Catalan despite widespread exposure. Thus, an active/passive distinction captures meaningful differences in sustained cognitive and linguistic engagement rather than mere exposure and has been repeatedly validated in prior Spanish-Catalan studies [16,28,29]. Moreover, in comparison to other bilingual populations, Spanish-Catalan bilinguals are relatively more homogenous with respect to immigration status and language distance variability.

The present study builds on this framework by examining whether bilingualism is associated with a later reported age at symptom onset and whether this association is consistent across clinical stages of typical AD, including cases with biomarker confirmation. By controlling for education, sex, and disease severity, and by anchoring analyses to both syndromic and etiologically confirmed AD diagnoses, this work aims to clarify bilingualism’s role as a cognitive reserve factor and to determine whether its protective association is evident irrespective of biomarker confirmation. We hypothesized that active bilingualism is associated with delayed symptom onset across clinical syndromes, serving as a cognitive reserve mechanism.

## 2. Methods

### 2.1. Study design and participants

This retrospective study analyzed data from the *Sant Pau Initiative on Neurodegeneration* (SPIN), a prospective cohort at the Memory Unit of Hospital Sant Pau, Barcelona, Spain [30,31]. Participants were drawn from a large, well-characterized cohort and met criteria for one of the following retrospective groups: (1) those meeting clinical criteria for amnestic dementia (*n* = 1,932) [30]; (2) those meeting clinical criteria for amnestic MCI (*n* = 1,443) [30]; and (3) a subset of groups 1 (dementia) and 2 (MCI), restricted to those demonstrating biomarker-confirmed AD (*n* = 1,068) [31]. Accordingly, individuals with clinical diagnoses of predominantly amnestic dementia or MCI were included regardless of biomarker availability, while a subset met etiological criteria for AD based on CSF Aβ42/Aβ40 ratio and phosphorylated tau-181, measured on the Lumipulse platform, applying previously established and validated Sant Pau cut-offs [30,31].

Individuals were excluded if they presented with a predominant behavioral, language, or motor clinical presentation, with syndromes consistent with atypical AD or other neurodegenerative disease phenotypes (e.g., corticobasal syndrome, Lewy body Dementia, amongst others), or bilingual profiles involving languages other than Spanish and Catalan, in order to limit linguistic and sociocultural heterogeneity. Language background was characterized based on self-reported current language use at the time of assessment. Participants were classified as active bilinguals if they reported actively using both Spanish and Catalan in their daily lives, or as passive bilinguals if they reported using Spanish actively while having passive exposure to Catalan. This classification, consistent with prior work in this population [16,27,29], was intended to capture functional bilingual language use, rather than formal assessments of language proficiency, balance, or age of acquisition.

In the biomarker-confirmed AD cohort, this classification resulted in 534 active bilinguals (315 women; mean age = 74.24 ± 6.48 years) and 534 passive bilinguals (341 women; mean age = 73.11 ± 6.56 years; see Table 1). Further, the distribution of active versus passive bilinguals in the amnestic dementia and MCI cohorts is also reported in Table 1. Across the three diagnostic cohorts, active bilinguals had significantly higher education than passive bilinguals, which was accounted for in the subsequent analyses. As all participants were recruited locally in Barcelona, the sample was relatively homogeneous with respect to immigration and cross-cultural differences, which are common confounders in prior bilingualism studies. All procedures were approved by the ethics committee of Hospital Sant Pau, and written informed consent was obtained from all participants in accordance with the Declaration of Helsinki.

**Table 1.** Descriptive characteristics by bilingual status and diagnostic cohort.

| Variable | Amnesic<br>Dementia:<br>Active<br>Bilingual<br>(*n* = 924) | Amnesic<br>Dementia:<br>Passive<br>Bilingual<br>(*n* = 1,008) | *p*-value | Amnesic<br>MCI:<br>Active<br>Bilingual<br>(*n* = 755) | Amnesic<br>MCI:<br>Passive<br>Bilingual<br>(*n* = 688) | *p*-value | Biomarker-confirmed<br>AD: Active<br>Bilingual<br>(*n* = 534) | Biomarker-confirmed<br>AD: Passive<br>Bilingual<br>(*n* = 534) | *p*-value |
| --- | --- | --- | --- | --- | --- | --- | --- | --- | --- |
| Women (%) | 531 (57%) | 626 (62%) | .042 | 401 (53%) | 410 (60%) | .015 | 315 (59%) | 341 (64%) | .116 |
| Age at first<br>evaluation (years) | 75.8 ± 7.23 | 74.9 ± 7.23 | .01 | 70.37 ± 9.7 | 70.11 ± 8.57 | .61 | 74.24 ± 6.48 | 73.11 ± 6.56 | .007 |
| GDS score at first<br>visit | 3.67 ± 0.65 | 3.65 ± 0.67 | .601 | 2.99 ± 0.42 | 3.02 ± 0.37 | .091 | 3.32 ± 0.61 | 3.36 ± 0.66 | .313 |
| Years of education | 10.8 ± 4.55 | 8.54 ± 4.5 | <.001 | 12.43 ± 4.67 | 9.94 ± 5.03 | <.001 | 11.46 ± 4.66 | 9.59 ± 4.91 | <.001 |

NOTE. Continuous data are mean ± standard deviation and categorical data are n (%). *p*-values reflect between-group comparisons using $\chi^2$ tests for categorical variables and independent-samples *t* tests for continuous variables, as appropriate. Abbreviations: MCI, mild cognitive impairment; AD, Alzheimer's disease; GDS, Global Deterioration Scale.

### 2.2. Demographic and clinical assessment

Demographic and clinical information was obtained from routinely collected data at the Memory Unit of Hospital Sant Pau. Demographic variables available for inclusion in this study consisted of age, sex, and years of formal education. Due to the objectives of the current study, reported clinical assessment measures include disease stage and reported age at symptom onset, rather than domain-specific neuropsychological performance. Age at symptom onset was defined as the chronological age at which participants and/or close informants first reported the emergence of symptoms, a standard approach in clinical dementia research [1,32]. Overall, disease stage was characterized using the Global Deterioration Scale (GDS) [33], which was recorded at the first clinical evaluation.

### 2.3. Statistical analysis

All statistical analyses were performed using *R* [34]. Descriptive statistics were first computed to summarize demographic and clinical characteristics across language groups and diagnostic cohorts. To examine the association between bilingualism and age at symptom onset, we fitted linear regression models via the car package [35], with age at symptom onset treated as a continuous dependent variable. Bilingual status (active vs. passive bilingualism) was included as the primary predictor of interest and modeled as a dichotomous variable. Separate regression models were estimated for each diagnostic cohort: (1) amnestic dementia, (2) amnestic MCI, and (3) biomarker-confirmed AD.

Covariates were included to control for potential confounding effects and to assess moderation. These covariates comprised years of formal education, sex, and GDS score at the first clinical visit. An interaction term between bilingual status and education was also incorporated to evaluate whether education moderated the association between bilingualism and age at symptom onset, as higher education has demonstrated potential protective effects [2]. Regression coefficients (β), corresponding *p*-values derived from the Wald tests, and 95% confidence intervals (CI) are reported for all models. Model assumptions including linearity, homoscedasticity, and normality of residuals were examined through visual inspection of diagnostic plots.

## 3. Results

### 3.1. Age of symptom onset in active vs. passive bilinguals in the amnestic dementia cohort

Linear regression analyses controlling for years of education, sex, and baseline GDS score revealed a significant association between active bilingualism and later age at symptom onset (*p* = .004). Bilingualism was associated with a delay of 1.42 years in symptom onset (see Table 2 and Figure 1). Estimated marginal means indicated an age at onset of 73.35 [CI: 72.68, 74.01] years for active bilinguals and 71.93 [CI: 71.24, 72.61] years for passive bilinguals. Greater years of education were significantly associated with earlier age at symptom onset (β = −0.37 per additional year of education; *p* < .001). The interaction between bilingual status and education (see Supplemental Table 1), as well as the main effect of sex were not significant.

**Figure 1.**
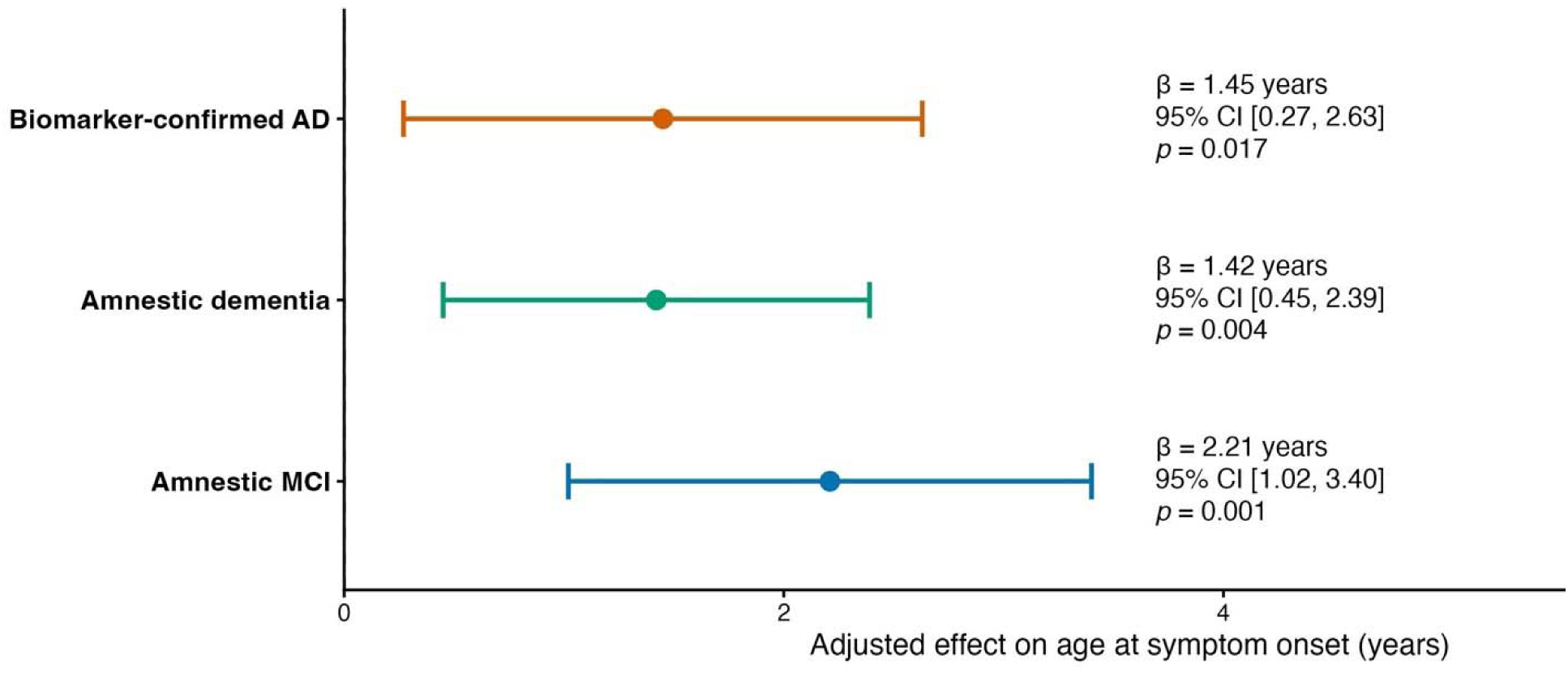
Active bilingualism is associated with later age at symptom onset across clinical stages. NOTE. Relative to passive bilinguals, active bilinguals had a delayed symptom onset across the amnestic MCI, amnestic dementia, and biomarker-confirmed AD cohorts. Adjusted effects are reported as β (active − passive bilingual) with 95% CI. Abbreviations: AD, Alzheimer’s disease; MCI, mild cognitive impairment; CI: Confidence Interval.

**Table 2.** Linear regression model for age at symptom onset in the amnestic dementia cohort (covariates: education, sex, and baseline GDS)

| **Predictor** | **$\beta$ (years)** | ***p*-value** | **95% CI** |
| --- | --- | --- | --- |
| Active bilingualism | 1.42 | .004 | 0.45, 2.39 |
| Education (in years) | −0.37 | < .001 | −0.48, −0.26 |
| Sex | 0.34 | .489 | −0.62, 1.3 |
| GDS (first visit) | 1.03 | .006 | 0.29, 1.77 |

NOTE. Abbreviations: GDS, Global Deterioration Scale; CI, Confidence Interval.

### 3.2. Age of symptom onset in active vs. passive bilinguals in the amnestic MCI cohort

Linear regression analyses revealed a significant association between active bilingualism and later age at symptom onset (*p* < .001). In this cohort, the main effect of active bilingualism consisted in a delay of 2.21 years (see Table 3 and Figure 1), when compared to passive bilinguals. Estimated mean age at onset was 69 years for active bilinguals and 66.78 years for passive bilinguals. Greater years of education were again significantly associated with earlier symptom onset (β = −0.49; *p* < .001). The interaction between bilingual status and education was significant, indicating that the bilingualism effect on symptom onset is significantly larger in less-educated participants. Sex was not a significant predictor.

**Table 3.** Linear regression model for age at symptom onset in the amnestic MCI cohort (covariates: education, sex, and baseline GDS)

| **Predictor** | **$\beta$** | ***p*-value** | **95% CI** |
| --- | --- | --- | --- |
| Active bilingualism | 2.21 | <.001 | 1.02, 3.4 |
| Education (in years) | −0.49 | <.001 | −0.61, −0.37 |
| Bilingualism × Education | −0.27 | .023 | −0.51, −0.04 |
| Sex | 0.33 | .588 | −0.86, 1.51 |
| GDS (first visit) | 0.41 | .572 | −1.01, 1.83 |

NOTE. Abbreviations: MCI, mild cognitive impairment; GDS, Global Deterioration Scale; CI, Confidence Interval.

### 3.3. Age of symptom onset in active vs. passive bilinguals in biomarker-confirmed AD cohort

Linear regression analyses demonstrated that active bilingualism remained significantly associated with delayed symptom onset even when restricting the sample to participants with biomarker-confirmed AD (*p* = .017). Active bilingualism was associated with a delay of 1.45 years (see Table 4 and Figure 1), when compared to passive bilingualism. Estimated mean age at onset was 72.03 [CI: 71.24, 72.82] years for active bilinguals and 70.58 [CI: 69.71, 71.45] years for passive bilinguals. Greater years of education again showed a significant negative association with age at onset (β = −0.27; *p* < .001; Figure 1). The interaction between bilingual status and education (see Supplemental Table 2), as well as the main effect of sex were not significant.

**Table 4.** Linear regression model for age at symptom onset in the biomarker-confirmed AD cohort (covariates: education, sex, and baseline GDS)

| **Predictor** | **$\beta$ (years)** | ***p*-value** | **95% CI** |
| --- | --- | --- | --- |
| Active bilingualism | 1.45 | .017 | 0.27, 2.63 |
| Education (in years) | −0.27 | <.001 | −0.39, −0.14 |
| Sex | 0.77 | .208 | −0.43, 1.97 |
| GDS (first visit) | −1.09 | .019 | −2, −0.18 |

NOTE. Abbreviations: AD: Alzheimer’s disease; GDS, Global Deterioration Scale; CI, Confidence Interval.

## 4. Discussion

Understanding why individuals with comparable AD pathology differ in the timing of clinical symptom onset is central to prevention and early intervention strategies [2,36]. Against this backdrop, we examined bilingualism as a potential source of variability in clinical expression and found that active bilingualism is associated with a later age at symptom onset across those presenting with predominant amnestic MCI, predominant amnestic dementia, and biomarker-confirmed AD. That is, active bilingual language use was linked to a later age at symptom onset across the clinical spectrum of typical AD. This association remained significant after controlling for education, sex, and disease severity, supporting the interpretation that bilingualism acts as a cognitive reserve factor rather than simply reflecting sociodemographic differences, considering that our cohort represents a relatively homogenous population relative to prior studies reporting such differences in bilingual cohorts. Importantly, the presence of this effect in biomarker-confirmed AD indicates that bilingualism delays symptom expression despite established AD pathology, strengthening a resilience-based interpretation and reducing concerns related to etiological misclassification in syndromic samples [25,32].

The magnitude of the bilingualism-associated delay ranged from approximately 1.4 years in amnestic dementia and biomarker-confirmed AD to just over 2 years in the amnestic MCI cohort, where the effect was most pronounced. This stage-dependent pattern is consistent with reserve and resilience models suggesting that compensatory mechanisms are most evident earlier in the disease course, when brain networks can still flexibly adapt to cope with the accumulating pathology [5]. Notably, these findings align closely with prior work from our group demonstrating that active bilinguals with biomarker-confirmed AD show better cognitive performance across multiple domains despite equivalent biological disease stage, providing direct evidence of cognitive resilience [27]. Further, this prior study demonstrated that active bilingualism was also associated with more favorable CSF and plasma biomarker profiles related to amyloid burden and neuroinflammation, suggesting the additional presence of resistance mechanisms. Taken together, these results support the view that bilingualism may contribute to both resilience and resistance processes, with the present study capturing their clinical manifestation as delayed symptom onset.

Previous retrospective studies and meta-analyses have reported larger delays, often in the range of four to five years, in bilingual individuals compared to their monolingual counterparts [17,25,26,37,38]. However, these studies relied on clinical diagnoses without biomarker confirmation. By anchoring analyses to etiologically confirmed AD and controlling for education and disease severity, the present study likely provides a more conservative and biologically grounded estimate of bilingualism’s protective association. In this context, a delay of one to two years should not be interpreted as a weaker effect, but rather as a reflection of bilingualism’s contribution once etiological heterogeneity and disease stage are explicitly accounted for.

Furthermore, the more modest magnitude of the bilingualism-associated delay observed here likely reflects key differences in sociolinguistic context and methodological rigor. Spanish-Catalan bilingualism typically involves early, lifelong, and societally balanced exposure to two closely related languages, often acquired implicitly and maintained across the lifespan, in contrast to more heterogeneous bilingual populations studied elsewhere that may include later language acquisition, immigration-related effects on cognition and brain health, or greater linguistic distance between languages. Additionally, in the current study, we compared active to passive bilinguals, whereas most prior studies had access to a “purer” monolingual comparison due to the distinct sociolinguistic contexts. These aforementioned contextual factors may amplify reserve and resilience effects in some cohorts, contributing to the larger magnitude of delays reported in the prior literature.

Education emerged as a robust covariate across cohorts, showing a paradoxical association with earlier reported age at symptom onset, particularly in amnestic MCI. Rather than indicating a detrimental effect of education, this pattern likely reflects earlier clinical detection and help-seeking behavior among more highly educated individuals, who may be more sensitive to subtle cognitive changes and more likely to seek evaluation at specialized memory clinics [39,40]. Taken together, these findings suggest that both education and bilingualism influence when symptoms are recognized and reported, rather than overall disease severity.

A strength of the present study is the relative homogeneity of the Spanish-Catalan bilingual context, which may help limit confounding influences associated with cross-national immigration or language distance, and allows a relatively more constrained examination of bilingual language use as a contributor to cognitive reserve and resilience. Moreover, the inclusion of biomarker-confirmed AD cases and adjustment for disease severity provide a more stringent test of bilingualism’s association with clinical expression than is possible in purely syndromic samples. Several limitations should be acknowledged. Focusing on a linguistically homogeneous population may limit generalizability to bilingual contexts characterized by later language acquisition, greater linguistic distance, or immigration-related cognitive demands, which may engage additional mechanisms relevant to cognitive reserve. Additionally, as participants were recruited from a specialized memory clinic, the sample may not be fully representative of the general population. Furthermore, the retrospective design precludes causal inference and relies on informant-reported age at symptom onset, which may be subject to recall bias, though this is a problem pervasive in the study of symptom onset more broadly.

In conclusion, this study provides robust and converging evidence that active bilingualism is associated with a delayed age at symptom onset across the clinical continuum of typical AD, including biomarker-confirmed cases. The protective association was most pronounced at the MCI stage and remained independent of education, sex, and disease severity. Together with prior work from our group demonstrating both resilience and resistance signatures in biomarker-confirmed AD [27], these findings reinforce bilingualism as a meaningful, lifelong experience shaping cognitive aging. Future studies integrating multi-site cohorts that represent the diversity of the bilingual experience more comprehensively, coupled with longitudinal biomarkers and neuroimaging, will be essential to further clarify the mechanisms through which bilingualism contributes to resilience and resistance in AD and related dementias.

## Data Availability

All data produced in the present study are available upon reasonable request to the authors.

## Acknowledgements

The authors would like to thank all participants and their families for their contribution to the research. We also thank all the clinical team members that were involved in the selection and assessment of participants, and the laboratory teams for sample handling, biomarker analyses, and structural support.

