## Supplemental Material for "Evidence for bilingualism as a cognitive reserve factor in biomarker-confirmed Alzheimer’s disease"

The interaction between bilingualism and education was not significant in the amnestic dementia and the biomarker-confirmed AD cohorts and therefore was not reported in the main manuscript. Results of these models including this interaction term are reported in the two tables below.

**Supplemental Table 1**. Linear regression model for age at symptom onset in the amnestic dementia cohort (covariates: education, sex, and baseline GDS)

| **Predictor** | **β** | ***p*-value** | **95% CI** |
| --- | --- | --- | --- |
| Active bilingualism | 1.42 | <.004 | 0.45, 2.38 |
| Education (in years) | −0.37 | <.001 | −0.48, −0.26 |
| Bilingualism × Education | −0.11 | **.313** | −0.33, 0.1 |
| Sex | 0.35 | .478 | −0.62,1.31 |
| GDS (first visit) | 1.03 | .007 | 0.29, 1.77 |

**Supplemental Table 2**. Linear regression model for age at symptom onset in the biomarker-confirmed AD Cohort (covariates: education, sex, and baseline GDS)

| **Predictor** | **β** | ***p*-value** | **95% CI** |
| --- | --- | --- | --- |
| Active bilingualism | 1.44 | .017 | 0.26, 2.63 |
| Education (in years) | −0.27 | <.001 | −0.39, −0.14 |
| Bilingualism × Education | −0.05 | **.693** | −0.29, 0.2 |
| Sex | 0.78 | .201 | −0.42, 1.98 |
| GDS (first visit) | −1.08 | .02 | −1.99, −0.17 |

Abbreviations: GDS, Global Deterioration Scale; CI, Confidence Interval; AD: Alzheimer’s disease.
